# Analytical Evaluation of Whole Genome Sequencing for Acute Myeloid Leukemia

**DOI:** 10.1101/2025.10.30.25339095

**Authors:** Weida Gong, Guidantonio Malagoli Tagliazucchi, Stewart Comer, Megha Ghildiyal, Mauro Chavez, Kan Nobuta, Tevfik Umut Dincer, Nandini Badarinarayan, Grace Kim, Quyen Bui, Carey Davis, Sean Truong, Severine Catreux, Taylor O’Connell, Colin Russell, Yutong Qiu, Francesco Brundu, Ashutosh Vashisht, Ashis Mondal, David Spencer, Sung Kim, Eileen de Feo, Ravindra Kolhe

## Abstract

Acute myeloid leukemia (AML) is the most common leukemia in adults and current methods rely on cytogenic and molecular profiling for AML classification and risk stratification. Recent advances have demonstrated that next generation sequencing (NGS) may improve prognostic prediction and risk stratification of AML patients. A tumor-only high coverage (∼220x) whole genome sequencing (WGS) method was developed and its analytical performance (limit of detection (LoD), sensitivity, and precision) was evaluated with clinical samples. Overall, the assay observed analytical sensitivity of 97.6%, 89.5% and 92.9% for small variants, Structural Variants (SV) and Copy Number Alterations (CNA), respectively comparing against reference sets from multiple modalities. LoD was evaluated as a function of sequence coverage and somatic variant allele frequency (VAF). At a given sequence depth of 140X, small variants (SNVs and indels) and SV achieved 95% detection rates when VAFs were 5% and 7.3%, respectively. CNA were detected with copy number fold change of 1.09 and 0.87 for duplication and deletion events, respectively. Further, loss of heterozygosity was detected with tumor purity of 17%. In Summary, the results demonstrate the WGS tumor-only (WGS TO) pipeline has high sensitivity in all variant types with a fast turnaround time of ∼5 days and can be used to identify variants indicative for AML treatment options.

## Introduction

Acute myeloid leukemia (AML) is the most common of all leukemias in adults and suffers from the lowest survival rate^1^. Although AML represents slightly less than 1% of all cancers, patients with AML were historically among the first cancer genomes sequenced under the Human Genome Project^2^ and it is one of the most genomically evaluated neoplasms. Currently, identification of genetic alterations in AML using cytogenic and molecular profiling are fundamental to diagnostic classification, risk assessment and therapy selection^3^.

Genomic studies have demonstrated that AML represents clonal evolution^4,5^ arising from an underlying leukemogenic process with a relatively small number of recurring mutations^5,6^. A small number of recurring mutated genes from hematopoietic stem cells undergo initial driver mutations and subsequent “subclonal expansion”^7,8^. These “hotspot” gene mutations are foundational to the current clinical diagnostic evaluation according to the World Health Organization (WHO) and other clinical guidelines for AML that rely on an array of cytogenic and molecular laboratory methodologies. Cytogenetic workups include conventional chromosomal banding analysis (CBA) and more recently optical genome mapping (OGM). Molecular workups to identify clinically relevant somatic variants include various modalities such as targeted and/or comprehensive genomic profiling (CGP) sequence panels, whole exome sequencing (WES), whole genomic sequencing (WGS), fluorescence in situ hybridization (FISH) and chromosomal microarrays (CMA). Recent advances in next generation sequencing (NGS), most notably WGS, have demonstrated clinical utility in characterizing clonal evolution in acute leukemias^4^.

Consequently, AML researchers have increasingly advocated the use of WGS since it is an unbiased genomic assay incorporating the broadest diversity of genomic alterations into a comprehensively singular and clinically relevant AML diagnostic classification schema^3^. This study reports on a tumor-only high depth coverage (∼220X) whole genome sequencing workflow (WGS TO) for AML genomic profiling with a rapid turnaround time of ∼5 days. This study aims to demonstrate the analytical performance, limit of detection (LoD), sensitivity, and inter/intra run repeatability to detect somatic single-nucleotide variants (SNV), insertions/deletions (indels), structural variants (SV) and copy number alterations (CNA) relevant to AML. The rationale for developing such higher depth sequencing coverage emanates from research that demonstrated higher depth coverage equates to higher detection rates of clinically relevant subclones with actionable therapeutic targets^5,6^. Thus, sequencing depth coverage > 140X allows the attainment of the key 5% somatic variant allele frequency (VAF) milestone^9^.

## Material and methods

### Overview

WGS TO is a tumor only whole genome sequencing workflow for the identification of somatic variants in AML samples. WGS TO workflow utilizes commercially available sample extraction protocols, Illumina PCR-free library preparation kit for wet-lab processing, sequencing on the Illumina Novaseq™ 6000 platform and DRAGEN™ Somatic (v4.2) Tumor Only analysis. The workflow was configured to target high sequencing depth (∼220x coverage), enabling high analytical sensitivity for variants with low VAF. The turnaround time for WGS TO from DNA extraction to reporting is ∼5 days. This includes wet lab procedures (<1 day), sequencing (∼2 days) and bioinformatic analyses (∼13 hours). (Figure 1 See Supplementary Material for details). 68 AML clinical cases, 46 Bone Marrow Mononuclear Cells (BMMC) and Peripheral Blood Mononuclear Cells (PBMC) samples from healthy donors, 3 cancer cell lines and Seraseq® myeloid mutation DNA mix reference material were used to assess the analytical performance of WGS TO. Specifically, these samples allowed the determination of analytical accuracy, reproducibility and analytical sensitivity (i.e. LoD).

**Figure 1.**
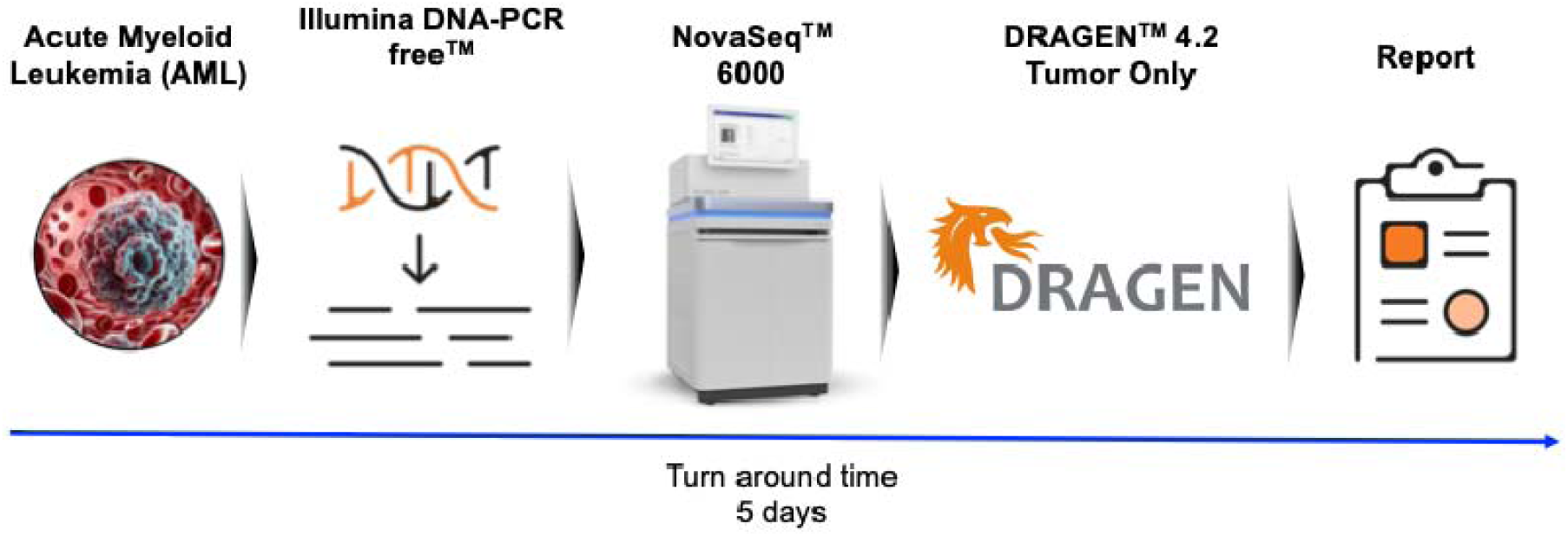
Summary steps of WGS TO. The WGS TO assay was implemented to process AML samples. The steps include DNA extraction, sequence library preparation, sequencing on a NovaSeq 6000 and sequence data analysis with DRAGEN Somatic in tumor only mode. The turnaround time (TAT) to process the samples is 5 days with sample preparation and analysis requiring <2 days.

### Clinical samples and cell lines

A total of 68 clinical samples obtained from 3 different clinical sources (23 samples from cohort A, 30 samples from cohort B and 15 samples from cohort C) were used to assess performance of the WGS TO assay. The diverse cohorts provide various comparator or orthogonal results which span from conventional methods (e.g. FISH and Karyotype) to sequencing-based methods (e.g. target gene panels) to detect variants that could be prognostic and/or diagnostic to AML. SVs and CNAs were characterized with both conventional methods (Karyotype and FISH) and newer techniques such as OGM and NGS. Sequencing-based methods and OGM have been reported to provide a higher resolution of genome wide genomic aberrations than standard of care. Having reference variant sets from multiple modalities enables the comparison of WGS TO to conventional and newer technologies. The reference variant list consists of 250 small variants, 38 SVs and 141 CNAs detected from multiple modalities. Refer to Supplemental Table 1 for a detailed list of variants from each modality. All patient samples were retrospective and consented for study research.

Analytical studies were supplemented with NA-12878, Kasumi-1, NOMO-1, HCC1187, and HCC1187-BL cell lines and the Seraseq® myeloid mutation DNA mix (also referred to as Seraseq samples). Kasumi-1 and NOMO-1 were selected for the presence of known structural variants (SVs), and HCC1187 replicates were used to determine the LoD of CNAs (See Supplementary Material and methods for additional details). Seraseq samples were also used to calculate the LoD of small variants.

### DNA extraction, Library preparation and Sequencing

DNA from cells lines, BMMC and PBMC were extracted using QIAgen AllPrep DNA/RNA Mini Kit (Qiagen, 80204). Input DNA concentrations were determined using the Quant-iT PicoGreen dsDNA Assay Kit (Thermo Fisher Scientific). DNA input was normalized to ∼350 ng in final volumes of 25 µL. Libraries were prepared using Illumina DNA PCR-Free Tagmentation Beads and Buffers (Illumina), Illumina DNA PCR-Free Purification Beads and Buffers (Illumina), and IDT for Illumina DNA/RNA UD Indexes Set A (Illumina). The library concentrations were quantified using KAPA SYBR dsDNA Q-PCR (Roche) and the libraries pooled in equal volumes at an ideal concentration of ∼1.5nM. The 4-plex library pools were then loaded and sequenced on an S4 flowcell with the NovaSeq 6000 platform. Preparation of the sequencer and sequencer-related consumables was done using standard Illumina workflow protocol.

### Sequence Data Analysis

Sequence alignment and variant calling were performed using DRAGEN Somatic v4.2 Tumor-Only^1^ in BaseSpace Sequencing Hub (BSSH) platform^2^ with hg38 as reference genome.

DRAGEN parameter settings were selected to optimize for AML samples by evaluating 46 healthy normal samples while incorporating hotspot variant information tailored to AML for DRAGEN analysis.

In addition, a custom filtering strategy was adopted to prioritize potentially clinically important variants for AML. Specifically, small variants were filtered using a list of 156 clinically important AML genes with coding consequences, SVs that overlap with a curated list of AML specific translocations and CNA larger than 5 MBs. A more detailed description of the variant prioritization procedure is provided in supplementary material.

### Analytical Sensitivity

A combined cohort of 68 AML samples were analyzed to assess analytical sensitivity. Variants detected by the workflow were compared against a reference set of variants from orthogonal methods including next-generation sequencing, FISH and karyotyping (Supplemental Table 1).

Sensitivity was computed as following:

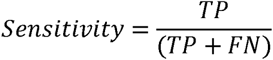

where TP represents variants detected by both the query set and reference set and FN being variants in reference set only. Variants below LoD (e.g. Small variants <5% VAF, CNAs < 500 kb) in the comparator set were not included.

### Analytical Precision

To examine analytical precision, clinical samples and contrived cell lines were sequenced in replicates within the same sequencing run or across different runs to assess repeatability and reproducibility in small variants, respectively. Table 1 describes samples, number of variants, and the precision testing scheme. Variants with VAF below LoD were excluded from the analysis resulting in the total variant count in Table 1.

**Table 1.** Analytical precision study setup. The table lists number of samples used, number of variants examined, number of intra-run concordance testing (repeatability) and number of inter-run concordance testing (reproducibility).

| Sample | # Samples | # Variants | # Repeatability | # Reproducibility |
| --- | --- | --- | --- | --- |
| Seraseq | 6 | 13 | 3 | 9 |
| Kasumi-1 | 10 | 41 | 12 | 33 |
| Clinical Samples | 15 | 11 | 3 | 6 |

### Limit of Detection

The LOD study was conducted to assess the impact of sequence coverage (C) in detecting small variant targets at a certain VAF with a 95% detection rate. Detection rate is defined as when DRAGEN generates an output which passes internal QC for the variant of interest. For example, variants may have read support for a specific mutation, however due to insufficient evidence or confidence, the variant is not reported. The following steps were performed to establish the LoD:

1. DNA from Seraseq samples was experimentally titrated with DNA from NA12878 to obtain different levels of tumor “purity” (0, 25%, 33%, 75%, 100%) (Supplemental Figure
2. with 5 replicates.
3. Each Seraseq titrated replicate was sequenced at nominal depth target 200X and then in silico down sampled by subsampling sequenced reads at different approximate coverage levels of 114x, 129x, 139x, 144x, 154x, 164x, and 189x.
4. At each down sampled coverage level, PROBIT (PROBability unIT) regression model was used to fit a curve of the detection rates (y-axis) onto the VAF (x-axis) of the variant of interest. (See Supplementary Methods for details).
5. VAF with 95% detection rate was estimated from the PROBIT curve at each downsampled coverage level.
6. A linear model was then fit between different down sampled coverage values and the respective VAF that resulted in a 95% detection rate. This fitted model was used to

interpolate the coverage required for 95% detection rate of variants at 5% VAF. This target VAF of 5% was selected of interest to provide high probabilities of identifying variants in lower abundant subclones with clinically actionable mutations.

The 95% detection rates for SV were evaluated at the sequence coverage set by the coverage that maintained a 95% detection rate for small variant VAFs as low as 5% for consistency.

NOMO-1 and Kasumi-1 cell lines were analyzed to conduct the SV LoD analysis. The cancer cell lines were titrated with NA12878 to create in-silico mixtures in 4 replicates at different level of tumor purities (0, 25%, 33%, 75%, 100%). PROBIT regression model was used to estimate detection rates at the down-sampled coverage of 140x for SV as was similarly done for small variants.

HCC1187 was used to conduct LoD analysis for CNA. In-silico admixtures were created by mixing FASTQ reads from HCC1187 with its matching normal HCC1187-BL. Each admixture pair was mixed at different proportions to target tumor purity levels at 5, 10, 20, 40 and 60%. Ten replicates were created for each admixture pair at each target purity level to estimate the 95% detection rate. PROBIT regression model was used to estimate the 95% detection rate at 140x (coverage set by the small variants LoD analysis) for each type of CNA (e.g. GAIN, LOSS and loss of heterozygosity (CNLOH)). The approach in determining the detectability of CNAs was similar to SVs and small variants, with the difference being that the fold change ratio of CNA was used instead of the VAF. CNLOH are copy number neutral events without changes in copy number, thus tumor purity, was used to determine the detectability at 140x coverage.

Supplementary Table 2 contains details of the list of variants examined for LoD. More details of the LoD analyses are also available in Supplemental Methods and Supplemental Figure 1.

## Results

### Analytical Sensitivity

Overall sensitivity was assessed from a total of 68 AML clinical samples. Sensitivity for small variant detection was 97.6% (n = 244/250, Figure 2a and Table 2), including 100% detection rate for 7 internal tandem duplications in FLT3 (FLT3-ITD). Estimated VAF from the WGS TO assay was highly correlated with VAF from the reference set (Supplemental Figure 2).

**Figure 2.**
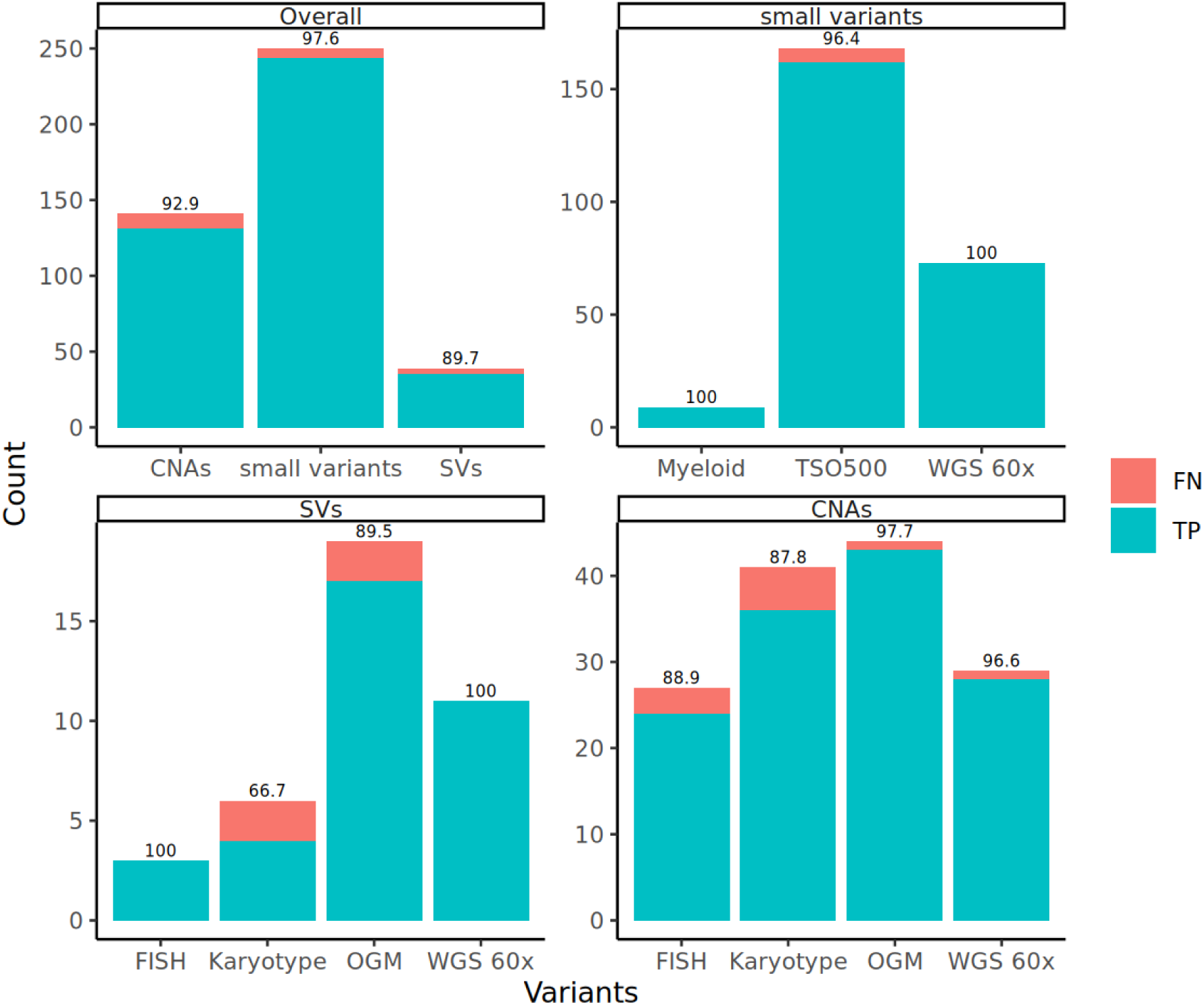
Sensitivity values of the WGS TO assay to detect different categories of variants across multiple modalities. a) Barplots with sensitivity of the variants from all 3 cohorts. b), c) and d) Highlight the sensitivity values to detect small variants, SVs, CNAs between different modalities. The y-axes represent the number of TPs (turquiose) and FNs (pink). The sensitivity values are reported on top of each bar. Variants below LoD (e.g. Small variants <5% VAF, CNAs < 500 kb) in the comparator set were not included.

**Figure 3.**
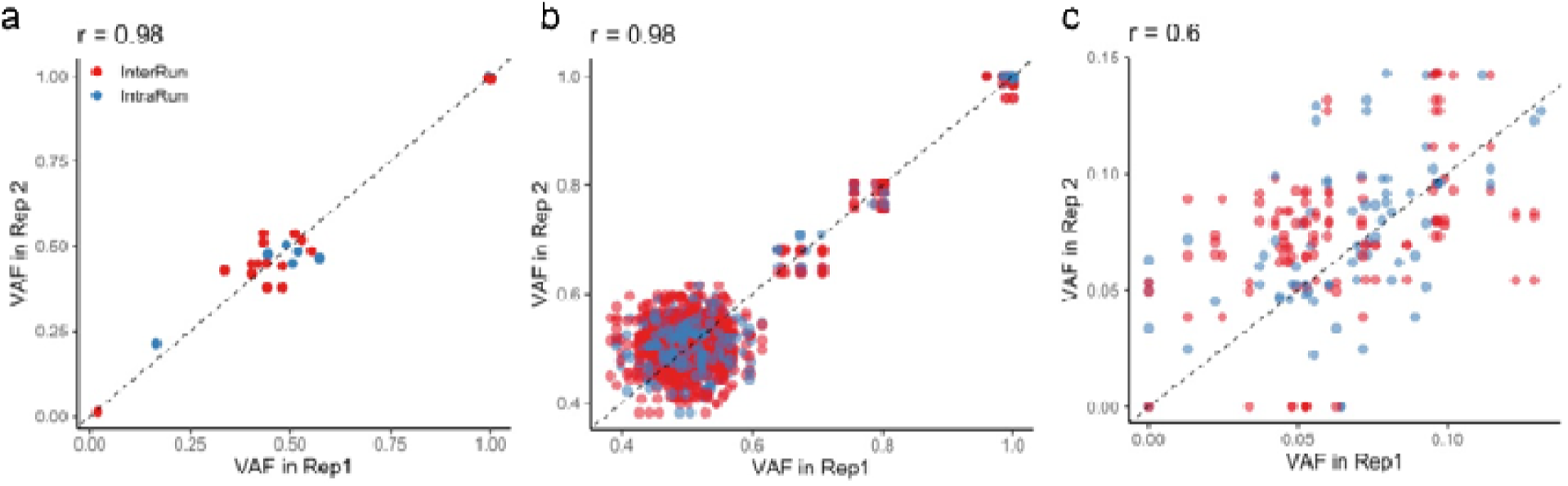
Inter- and intra-run performance. Correlation of variant allele frequency (VAF) of variants between replicates in a) DLS samples, b) Kasumi-1 and c) Seraseq. Pearson correlation coefficient are indicated on top of each sub-plot

**Table 2.**
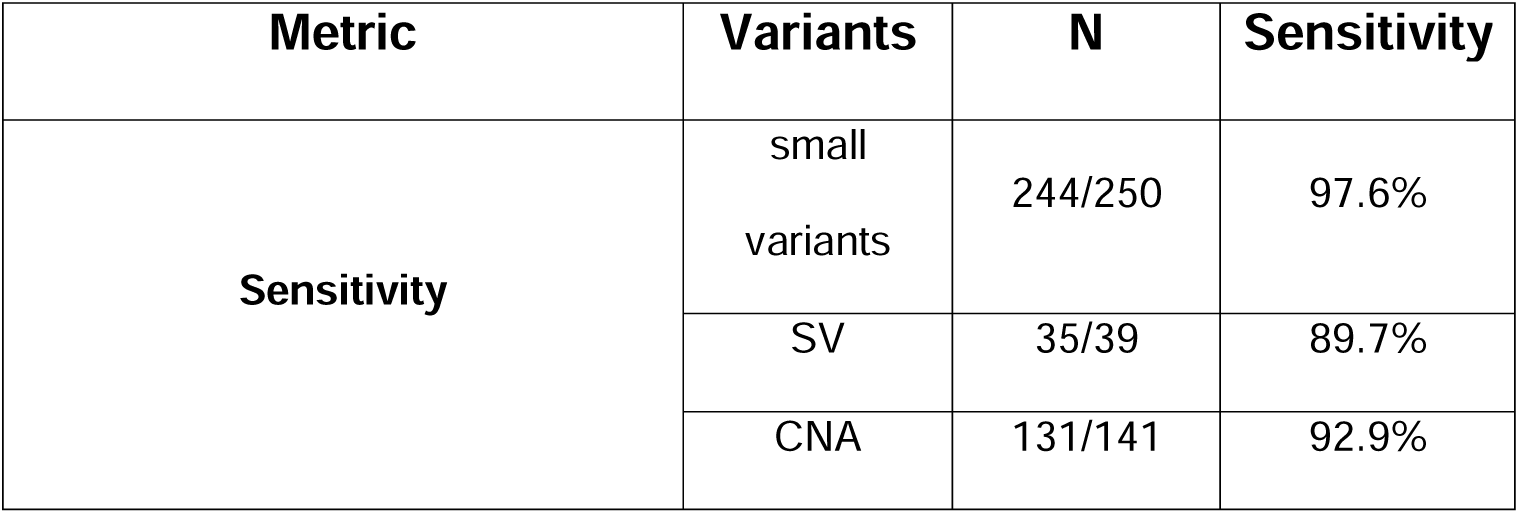
Sensitivity of the WGS TO assay. Sensitivity was measured against a total of 68 AML samples. Sensitivity is reported for each variant type. Variants below LoD (e.g. Small variants <5% VAF, CNAs < 500 kb) in the comparator set were not included.

Sensitivity was 83.9% (34/38) and 92.9% (131/141) for SVs and CNAs (Figure 2a, Table 2), respectively.

### Benchmark sensitivity against multiple modalities

Sensitivity was assessed in AML samples from 3 different cohorts where the comparator set of variants were measured using different technologies, including both traditional methods (i.e. karyotyping and FISH) and sequencing-based methods. Here, this study further expands on each modality to benchmark performance of the WGS TO against multiple technologies.

Small variants in the comparator set were detected using 60x whole genome sequencing (WGS) in cohort A and target panel sequencing (TruSight Oncology 500 Assay, TSO500 and/or

TruSight Myeloid Sequencing Panel, myeloid panel) in the other 2 cohorts. The WGS TO detected 73/73 small variants comparing with 60x WGS whereas WGS TO detected 162/168 and 9/9 small variants comparing with TSO500 and myeloid panel, respectively (Figure 2b). The 6 variants not reported by WGS TO are summarized as follows (Supplemental Table 1 and Supplemental Table 3):

1. 1 variant (FAM227B: c.574+38203A>G) was detected by WGS TO but annotated as a possible germline variant and filtered out
2. 1 variant (PMS2: p.L729fs*6) was missed but was detected by WGS TO in corresponding pseudogene
3. 1 variant (ICOSLG: c.817+398G>A) was detected but filtered out due to low quality
4. 3 variants (NRG1: c.37+649G>C; RANBP2: p.L554L; RAD21: c.764T>A) were not

reported by WGS-TO due to very low read support.

For SVs, WGS TO detected all 11 SVs compared with 60x WGS. WGS TO reported 17/19 SVs when compared against OGM (Figure 2c, Supplemental Table 3) with very few sequence read support for the 2 missed SVs under IGV inspection (Supplemental Figure 3-4). WGS TO detected 4 of 6 SVs and 3 of 3 SVs comparing with Karyotype and FISH, respectively. Similar to OGM, IGV inspection of the missed variants demonstrated no sequence read support for the 2 SVs specifically reported by karyotyping (Supplemental Table 3).

For CNAs, WGS TO detected 24/27 (88.9%), 36/41 (87.8%), 43/44 (97.7%), 28/29 (96.6%) events when compared with FISH, Karyotype, OGM, and WGS 60x, respectively (Table 3, Figure 2d). Overall, CNAs were reproducible across different modalities, however there are marked differences with increased concordance as resolution of the modality increases. In the case of the discordant result between WGS TO and WGS 60x, the SV reported by WGS 60x was noted as a low confidence variant in the reference dataset.

**Table 3.** Sensitivity of the WGS TO assay compared against multiple modalities. A total of 68 AML samples were analyzed. Sensitivity is reported for each variant type by orthogonal method.

| Metric | Type of variants | Orthogonal technology | N | Performance |
| --- | --- | --- | --- | --- |
| Sensitivity | small variants | TSO | 162/168 | 96.4% |
|  |  | WGS | 73/73 | 100% |
|  |  | Myeloid | 9/9 | 100% |
|  | SV | WGS | 11/11 | 100% |
|  |  | OGM | 17/19 | 89.5% |
|  |  | Karyotype | 4/6 | 66.7% |
|  |  | FISH | 3/3 | 100% |
|  | CNA | OGM | 43/44 | 97.7% |
|  |  | WGS 60x | 28/29 | 96.6% |
|  |  | Karyotype | 36/41 | 87.8% |
|  |  | FISH | 24/27 | 88.9% |

For a subset of the samples, multiple orthogonal testing results were available. This not only provides an opportunity to compare WGS TO against each modality, but also enables the comparison of variants across different reference standards. There was consistently high concordance between WGS-TO and OGM (97.7%), however WGS TO was slightly lower in concordance against other conventional lower resolution methods (Karyotype and FISH). WGS TO failed to detect 5 CNAs compared with Karyotype. Of note, these CNA’s were isolated to Karyotyping results and thus also not reported by OGM or FISH. When comparing WGS TO against FISH, WGS TO failed to detect 3 CNAs, however, interestingly, all of these CNAs were also not reported from other orthogonal methods. In one example, FISH reported both a gain and a loss of 5p (CohortC_232) whereas it was not reported by other methods (Supplemental Table 1). Overall, a total of 10 CNAs were missed in WGS TO comparing against OGM, FISH and Karyotype and none of these CNAs had consistent supporting evidence across the different modalities. This suggests further experimentation and analysis are warranted to assess potential root causes of the differences in results.

In summary, WGS TO achieve an overall sensitivity of 97.6%, 89.7% and 92.9% for small variants, SVs and CNAs comparing against a set of reference methods. A total of 6 small variants, 4 SVs and 10 CNAs were reported by reference methods but not reported by WGS TO. These variants upon further inspected under IGV showed very few if any sequence read support the variant of interest (minus one small variant where it was detected by WGS TO but filtered out as a possible germline variant. Supplemental Table 3). In addition, the 4 SVs and 10 CNAs not reported by WGS TO had inconsistent results based on other reference methods.

Therefore, further additional testing would help to confirm and provide more accurate estimates of performance of WGS TO.

### Analytical precision

Fifteen clinical AML samples were sequenced in replicates within the same flowcell (intra-run) and across different flowcells (inter-run) to assess repeatability and reproducibility of small variants. WGS TO achieved a concordance of 100% for inter and intra run comparison (Table 4). Further, there was a statistically significant positive correlation in VAF between replicates (Figure 4).

**Figure 4.**
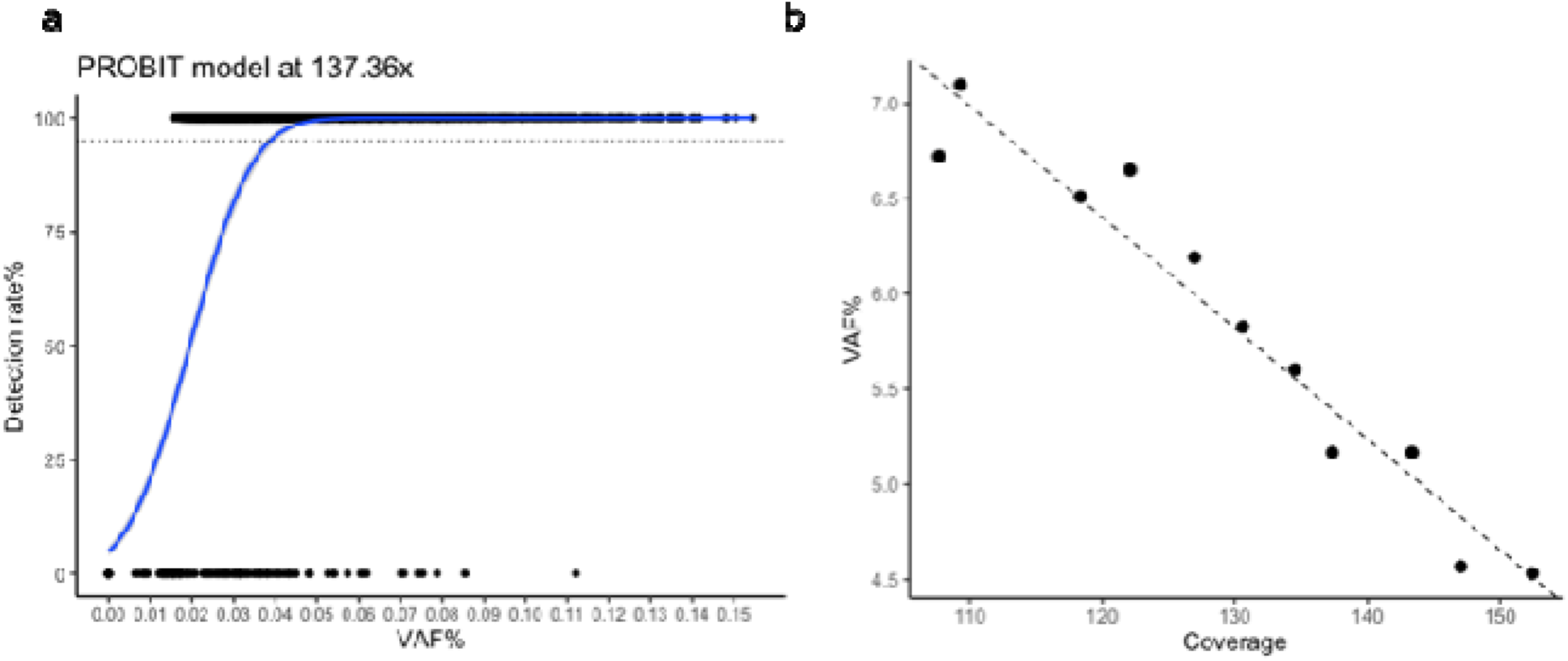
Limit of detection of the WGS TO assay. a) PROBIT regression model, with expected VAF (y-axis) at 140x of coverage. b) Results of the LoD analysis and linear regression between coverage (x-axis) and VAF (y-axis). A minimum coverage of 140X was determined from the LoD analyses of small variants

**Table 4.** Analytical precision of the WGS TO assay. Listed are median of repeatability and reproducibility for each sample. Numbers in parathesis denotes the minimum and maximum of concordance rate for each sample.

| Sample | Metric |  |
| --- | --- | --- |
|  | Repeatability | Reproducibility |
| Seraseq | 92% (85% - 100%) | 92% (85% - 100%) |
| Kasumi-1 | 100% (100% - 100%) | 100% (100% - 100%) |
| DLS | 100% (100% - 100%) | 100% (100% - 100%) |

Furthermore, 6 replicates of Seraseq and 10 replicates Kasumi-1 cell lines were sequenced to assess analytical precision of the WGS TO assay in cell lines. For Seraseq, concordance was computed between replicates using 13 small variants in the reference set and WGS TO reported a median concordance rate of 92.3% for repeatability and reproducibility, respectively (Table 4). There are 2 discordant variants across replicates (AXSL1: c.1934dup and ASXL1: c.1900_1922del). The two variants were reported in 4 reps but were filtered out in the other 2 reps due to low quality filters and/or too few supporting reads. The observed VAF for the two variants are around the LoD of the WGS TO assay (5.43% and 4.99%, respectively.

Supplemental Table 4), which could account for the inconsistent detection across replicates. 41 small variants were analyzed to compute concordance rate between replicates for Kasumi-1.

Repeatability and reproducibility were both reported at 100% (Table 4), highlighting a high concordance in small variant calling performance in the WGS TO assay. A full list of variants assessed in analytical precision is provided in Supplemental Table 4.

### Limit of Detection

The LoD study evaluated the impact of sequence coverage and the VAF of small variants that resulted in 95% detection rates. Seraseq was titrated with NA-12878 in replicates to create a series of admixture samples with small variants with known VAF that ranged from 0.63% to 13.06% (Supplemental Table 2 for experiment setup). The *in silico* down sampled Seraseq admixtures at specific sequence coverage allowed the PROBIT modelling of detection rates as a function of VAF%. That is, for each sequence coverage, the PROBIT model determined the VAF % that results in a 95% detection rate (Fig 4a). A linear model was then fit between coverage and the PROBIT determined VAF % to interpolate the sequence coverage requirements in detecting small variants with ∼ 5% VAF (Refer to Method section for more details). Figure 4b highlights this relationship of coverage (x axis) required to achieve a 95% detection rate for small variants at certain VAF (y axis) where higher coverage correlates to consistent detection of variants with low VAF. The coverage was estimated to be 140x to reach a detection rate of 95% for small variants at or above 5% VAF.

For SV, detection limits were evaluated at 140x for consistency. NOMO-1 and Kasumi-1 were experimentally titrated with NA-12878 to create a series of cell line admixtures for SVs with known VAF (SV VAF ranges from 1.75% - 27.60%, Supplemental Table 2 for experiment setup). The titrated series were then sequenced at nominal depth prior to in silico down sampling. A PROBIT model on the in silico down sampled 140x titrated data was fit to estimate the LoD for SV. The analysis demonstrated that SV with 7.3% VAF can be detected with 95% probability at 140x coverage (Table 5).

**Table 5.**
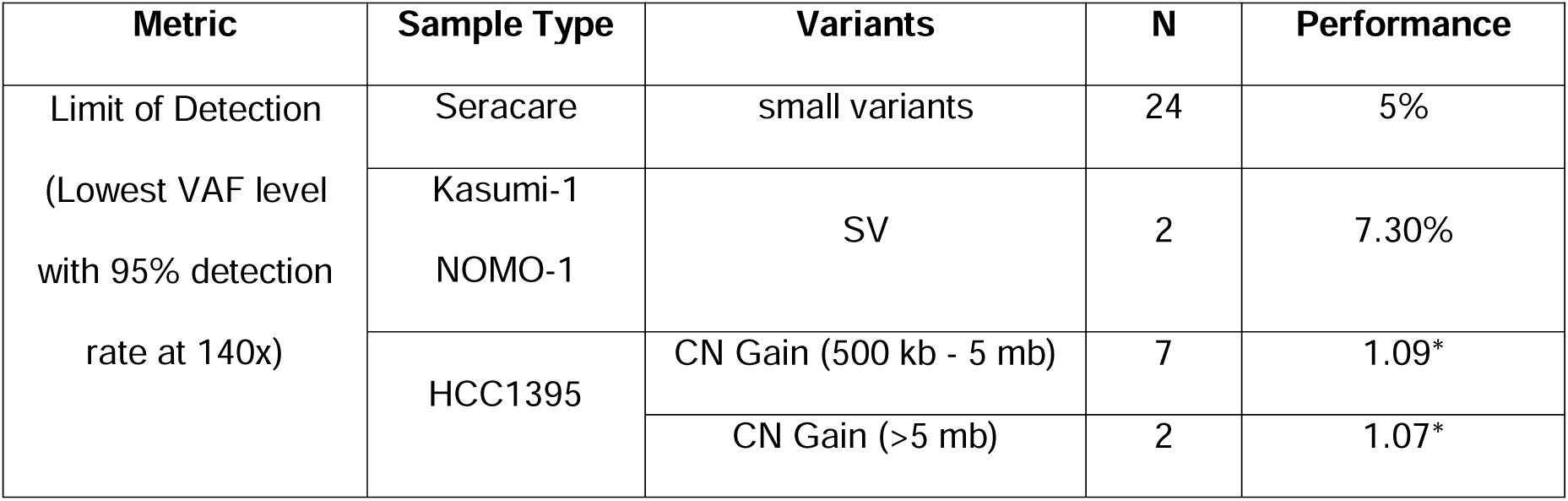
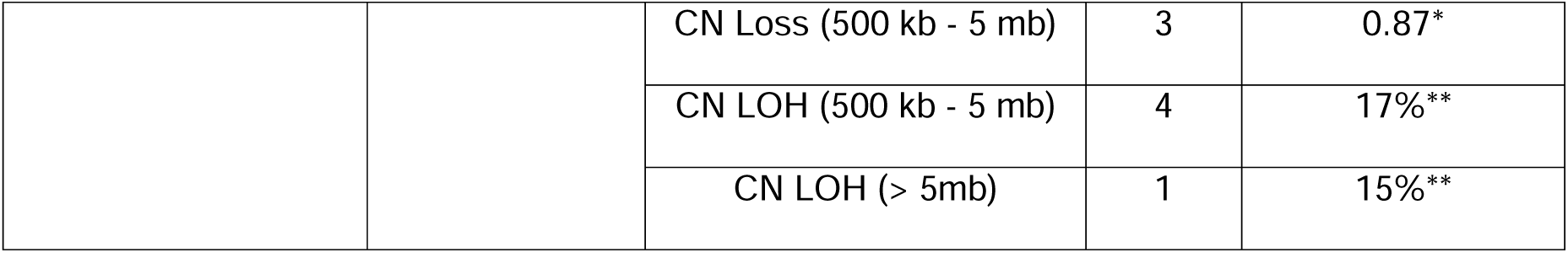
LoD of the WGS TO assay. Established LoD at 140x coverage for each variant type. N represents number of variants examined in each variant type. LoD for small variants and SVs were reported in VAF. CN fold change (*) or tumor fraction (**) were reported for CNAs.

For CNAs, HCC1187 was used to evaluate detection rates and, again, analysis was focused on coverage at 140x for consistency with small variant detection rates. The computation of the detection rates for CNA requires cell lines to be titrated with matched normal genomic backgrounds as DRAGEN leverages expected allele frequency ranges in the algorithm that determines CNVs. HCC1187 contains multiple CNAs and have a matched normal genomic background to allow for experimental titrations to evaluate the analytical sensitivity in detecting CNAs. In addition, the ability to detect CNAs in cancer is assumed to be similar regardless of cancer type, i.e. CNA detection in breast cancer is expected to perform similarly to CNA detection for hematological cancers.

In-silico admixtures were created with HCC1187 and its matching normal HCC1187-BL to create a series of samples with known tumor fraction. Multiple replicates were created at each target tumor fraction (Supplemental Table 2). CNA events were stratified by CNA type (Gain, Loss and LOH) and size of the event (500KB to 5 MB and > 5 MB) and established performance for each CNA type and size group. The detection limits were 1.09- and 0.87-fold change for CN Gain and Loss events, respectively, when coverage is set at 140x. In addition, the CNLOH’s detection limit, measured as tumor purity, was 17% (Table 5).

## Discussion

AML is one of the most genomically evaluated neoplasms, which has generated a deep compendium of research. The last two decades have uncovered critical insights linking genomic alterations to specific AML subtypes and therapeutic regimens that match selective prognostic risks. Consequently, diagnostic laboratory testing is an essential modality to interrogate the cytogenetic and molecular heterogeneity in AML samples. The comprehensive characterization of these molecular features is crucial for therapy selection as well as providing prognostic risk stratification under current ELN guidelines^10^ and Myelodysplastic Syndromes to AML critical diagnostic cutoff criteria under WHO and ICC guidelines^11^. A variety of techniques spanning the decades from the 1960s have been employed to detect these molecular abnormalities in AML including CBA, FISH, CMA, PCR, and sequencing starting with Sanger techniques and evolving into NGS with CGP, WES and WGS. Ultimately, any technological modality needed to effectively characterize AML requires the ability to detect SNV, insertions, deletions, CNA and SV alterations, the latter two of varying lengths and complexity. All modalities are required to demonstrate sufficient resolution to meet acceptable detection sensitivity.

Within the last decade, two laboratory methodologies are being recognized for their effectiveness in detecting the increasing breadth of genomic alterations. The methods used for cytogenetic evaluations, particularly complex SVs, has evolved from CBA in the early 1960s to OGM. OGM was first described in 1993 but demonstrated limited clinical utility due to high error rates and low throughput^12^. Technologic advances in microfluidics, enzymology, image automation and data analytics have propelled OGM into consideration as a more recent standard of care testing modality for AML, where diagnostic guidelines are increasingly defined genomically. OGM, with a resolution > 500 bp and 300-400x coverage depth^13,14^, can detect 5- 10 Mb structural anomalies and provides effective discrimination with highly repetitive chromosomal regions including segmental duplications and complex genomic rearrangements. OGM, however, lacks capability for base pair level resolution, which is essential for the effective detection of SNV and indel variants required for the accurate diagnosis of AML. This requires higher resolution techniques such as sequencing. The utility of WGS as a potential NGS option stems from similar advantages shared with OGM, namely an unbiased method with whole genome wide detection capability. Unlike “hot-spot” PCR, targeted CGP or WES that do not allow for the comprehensive detection throughout the genome, WGS allows both exonic and intronic variant detection. This capability is important for AML given the significant number of recurrently mutated variants that have been recognized as being intergenic^3^.

In this study, a WGS assay (WGS TO) is presented which features fast-sequencing and efficient library preparation, high depth coverage (> 140x) for enhanced analytical sensitivity for somatic variants with VAF ≥ 5% and standardized bioinformatic pipeline using DRAGEN. Optimization in wet lab procedures as well as leveraging the computational power of DRAGEN enables a total turnaround time of 5 days from DNA extraction to VCF report, which is on par with PCR-based molecular assays^15^. Analytical performance of the WGS TO assay was evaluated using clinical samples and contrived cell lines with reference set from multiple modalities. Overall, analytical sensitivities were 97.6%, 89.5% and 92.9% for small variants, SV and CNA respectively. In addition, the WGS TO assay achieved a 100% (7/7) detection rate of internal tandem duplications in FLT3 (FLT3-ITD). The missed variants were further inspected under IGV and there was few if any sequence reads that supported the variants. WGS TO performed as expected and did not report out the variants as there was not enough evidence when VAF < 2%. The difference in VAF from WGS TO and the reference method could be attributed to inherent biological variability between the samples or inherent limitations when comparing different assays with different biases and resolution. Furthermore, SVs and CNAs that resulted in reduction in performance typically had inconsistent results across different modalities in the reference set. Therefore, further experimentation effort is warranted to disentangle conflicting results observed in this study.

Analytical performance of the WGS TO was benchmarked against a reference variant set from both conventional methods (i.e. FISH, Karyotype) and newer techniques (sequencing and OGM). Consistent with literature, analytical sensitivity was higher when compared WGS TO with OGM/sequencing methods than conventional methods (Table 3). Interestingly, WGS TO reported a total of 339 additional small variants with coding consequences, 32 additional SVs and 122 additional CNAs that are larger than 5 MBs which were not detected by the orthogonal methods in the reference set (Supplemental Table 5). The SVs and CNAs reported only by WGS TO are potentially prognostic to AML and are informative for treatment decisions. The additional variants highlight the sensitive resolution of high depth WGS TO assay and potentially provides a more comprehensive characterization of variants across the whole genome.

However, additional validation work is warranted to confirm the variants identified by WGS TO.

## Conclusions

WGS TO assay is a fast and efficient workflow that comprehensively profiles SNV, indels, CNA, and SV with high sensitivity. This assay features rapid sample processing, improved hand on time, and simplified bioinformatic processing and run-time. This unbiased WGS assay provides enhanced analytical sensitivity comparable to other targeted and exon-based NGS genomic sequencing methods. Additionally, highly repetitive sequence variants, reportedly a limitation for NGS technology^16^, were effectively detected, exemplified by the 100% concordance in detecting FLT3-ITD. Overcoming previously described limitations of NGS technology, high-depth WGS may serve as an effective single test modality to a larger number of AML patients, on the order of 94% based on some estimates^3^, that may not need additional or confirmatory findings to create a final clinical report. Analytical performance demonstrates that this tumor-only WGS approach provides high accuracy in identifying variants relevant for AML. AML is a disease of a small number of recurrently mutated genes initiated from hematopoietic stem cells with initial driver mutations undergoing clonal expansion creating “founding” clones leading to “subclones” through “cooperating driver” mutations. The higher depth coverage allows for increased detection of AML subclones that might not otherwise be detected with lower coverage depth or other targeted assays, but may be clinically actionable^5,6,8^. This potentially creates an avenue for further investigations leading to improved and highly sensitive risk stratification schema.

## Data Availability

Data produced in the present study may be available upon reasonable request to the authors based on the approved IRB guidelines

## Acknowledgments

The authors thank Josh Bernd, Danielle Collins and Teng Vang (Illimina Inc.) for assistance in generation of sequencing data, Jennifer Becq, Kimberly Gietzen, Theo Heyns, Martina Mijuskovic, Lisa Murray, Alina Ozuna, Erfan Sayyari and Konrad Scheffler (Illumina Inc.) for assistance in sequencing data analysis and variant interpretation.

## Footnotes

1 Available in DRAGEN v4.4: https://help.dragen.illumina.com/product-guides/dragen-v4.4/dragen-apps/dragen-heme-wgs-to-pipeline

2 https://www.illumina.com/products/by-type/informatics-products/basespace-sequence-hub.html

