## Supplemental Material for "Analytical Evaluation of Whole Genome Sequencing for Acute Myeloid Leukemia"

### Supplementary methods

#### Data analysis and visualization

Visualization and analyses of primary sequencing metrics was conducted with JMP Statistical Discovery software. All JMP analyses utilized data generated from DRAGEN 4.2. To investigate secondary metrics, custom R and Python scripts were used.

#### Systematic noise file

Systematic Noise files (SNFs) were created to filter out sequencing artefacts/noises. The SNF was created from a cohort of 42 and 43 normal samples for SNVs and SVs, respectivelyß.

#### AML specific hotspot file

DRAGEN allows the usage of hotspot VCF files with the positions of regions where more somatic mutations are expected for boosted sensitivity. All the variants in one of these positions are also tagged as hotspot. In this work, an AML SNV hotspot VCF file was generated aggregating data from different publicly available dataset of tumor normal analysis[1–3]. The default SV hotspot VCF file implemented in DRAGEN was used.

#### Variant filtering system

A custom pipeline was implemented and used to prioritize AML specific variants (SNVs and SVs), based on their somatic status, predicted consequence, overlap with hotspot regions, external files, with a clinical interest.

Specifically, the following filtering procedures are applied for each variant type:

1. Small Variants:
    a. Variants annotated as somatic or tagged as hotspot.
    b. Variants with deleterious coding consequence (e.g. Missense variant, Frameshift variant).
    c. Overlap with 156 AML gene list.
2. Structural variants:

a. Size > 100k bp for SVs that are not translocations or inversions.
b. Overlap with 613 recurrent AML SVs list.

Copy number alterations are also prioritized with these definitions:

a. Size > 5 MB.

Variants after the filtering were compared against the reference variant set. Variants that were not in the reference variant set were reported as additional variants from WGS TO (Supplemental Table 5).

**<u>LoD of small variants and structural variants</u>**

SNVs were surveyed via Seraseq samples with known small variants which included SNVs, indels, and ITDs. NPM1 and ASXL1:c1900_1922del were not detected at full coverage (~200x) and were not considered for LoD calculation. Structural variants were surveyed via NOMO-1 and Kasumi-1 samples each with a known translocation. Each of these cell lines were mixed with NA12878 to target different VAFs. The backbone of the titration is relative to the reference genome and the study aims to evaluate the effect of reducing the alternate allele relative to the reference base. However, admixtures with different germline profiles could potentially produce FP calls at the sites with germline mutations. The FP calls may introduce noise in INDEL error rate estimation so it may underestimate the sensitivity for INDELs. Therefore, the LoD measured here can serve as a conservative estimate.

For small variants, cell line admixtures were down-sampled at multiple depth (Supplemental Figure 1) to estimate the minimum coverage required for 95% detection rate of small variants > 5% VAF.

For SVs, LoD was evaluated and established at the sequence coverage set by the minimum coverage that maintains a 95% detection rate for VAFs as low as 5% for small variants for consistency. PROBIT regression model was used to estimate LoD at down-sampled coverage of 140x for SV.

**LoD of copy number alterations (CNAs)**

For CNAs, HCC1187 was reviewed and selected for CNA LoD establishment. Although HCC1187 is not specific to hematological cancers, this cell line contains many CNAs that can be used to infer generic CNA analytical sensitivities for the assay that should still be applicable for hematological cancers. In-silico admixtures were created by mixing FASTQ reads from HCC1187 with its matching normal HCC1187-BL. Each admixture pair was mixed at different proportions to target tumor purity levels at 5, 10, 20, 40 and 60%. Ten replicates were created for each admixture pair at each target purity level to estimate LoD. Similarly with small variants and SV, PROBIT regression model was used to estimate LoD at 140x for each type of CNA (e.g. GAIN, LOSS and loss of heterozygosity (CNLOH)).

DRAGEN Somatic analyses were launched for each tumor-normal admixture. Variants from VCF files were compared with a list of CNA candidates to compute LoD. Only CN events with "PASS" label in VCF file and with at least 70% overlap in length with corresponding CN event from the candidate list are considered TPs. CN events were categorized into two groups by size: 500 KB to 5 MB and > 5 MB. LoD analyses were stratified by CN type (CN gain, CN loss, and CNLOH) and size. For each CN category, a PROBIT regression model was fitted where tumor purity was used as explanatory variable to predict detection rate. A point estimate of tumor purity and fold change from the PROBIT regression model was provided as an estimate of LoD with 95% detection rate.

**Assessing reproducibility and repeatability using contrived material**

The levels of concordance of the Seraseq samples, analyzed at full-depth, were computed for repeatability and reproducibility. Repeatability was measured by performing a pair-wise comparison of the samples in the same run. To compute the reproducibility, a pair-wise comparison of the samples between two different runs was performed. In both comparisons, as a proxy for concordance between two samples, we computed the TP/[TP+FP+FN]. The Kasumi-1 reference set consisted of 41 somatic small variants (SNVs and INDELs, Supplemental Table 4). The levels of concordance of the Kasumi-1 samples at full-depth were computed in the same fashion as for SeraCare for repeatability and reproducibility.

#### Supplemental Figures

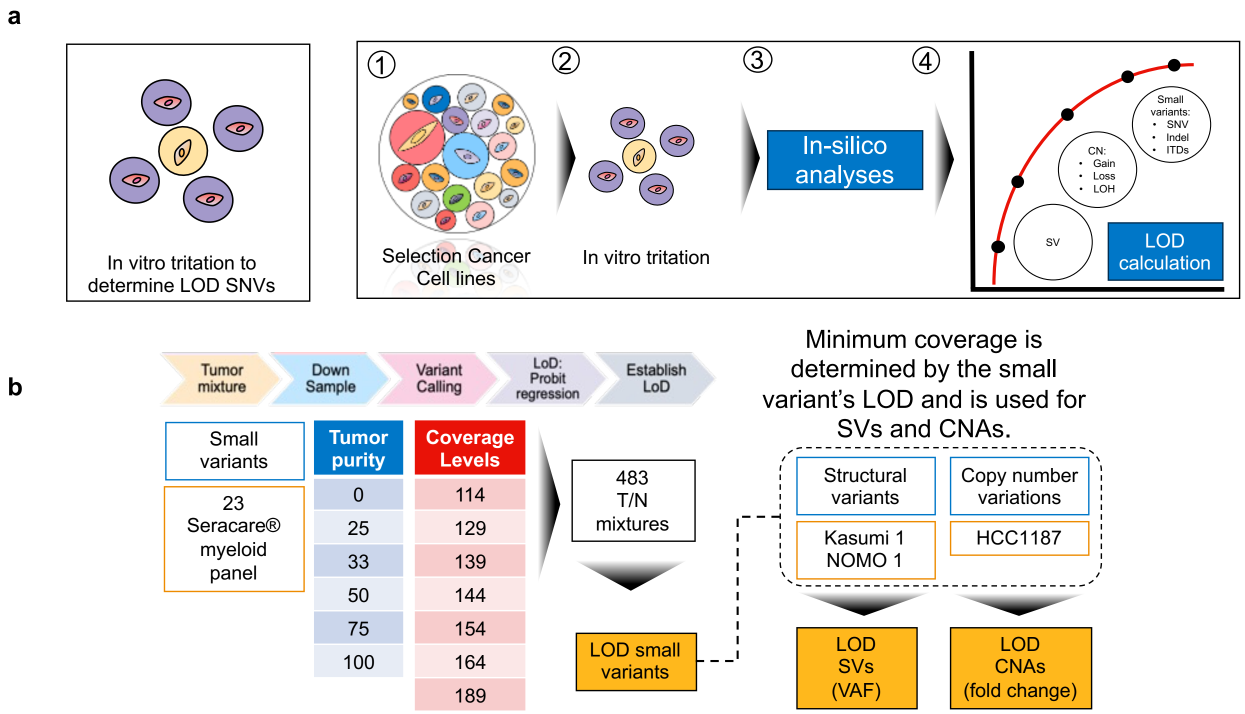

**Supplemental Figure 1. Computational framework to establish LoD.**

a) The computation of the analytical sensitivity was based on in-vitro titration and in-silico approaches. A pool of selected cancer cell lines was mixed with NA12828 to target different level of tumor purity to assess the LoD b) SeraCare cancer lines were used to establish the LoD of the small variants. For small variants, LoD was determined as the minimum coverage required for 95% detection rate of small variants > 5% VAF. For SVs and CNAs, LoD was then determined as the minimum VAF and tumor purity with > 95% detection rate at 140x.

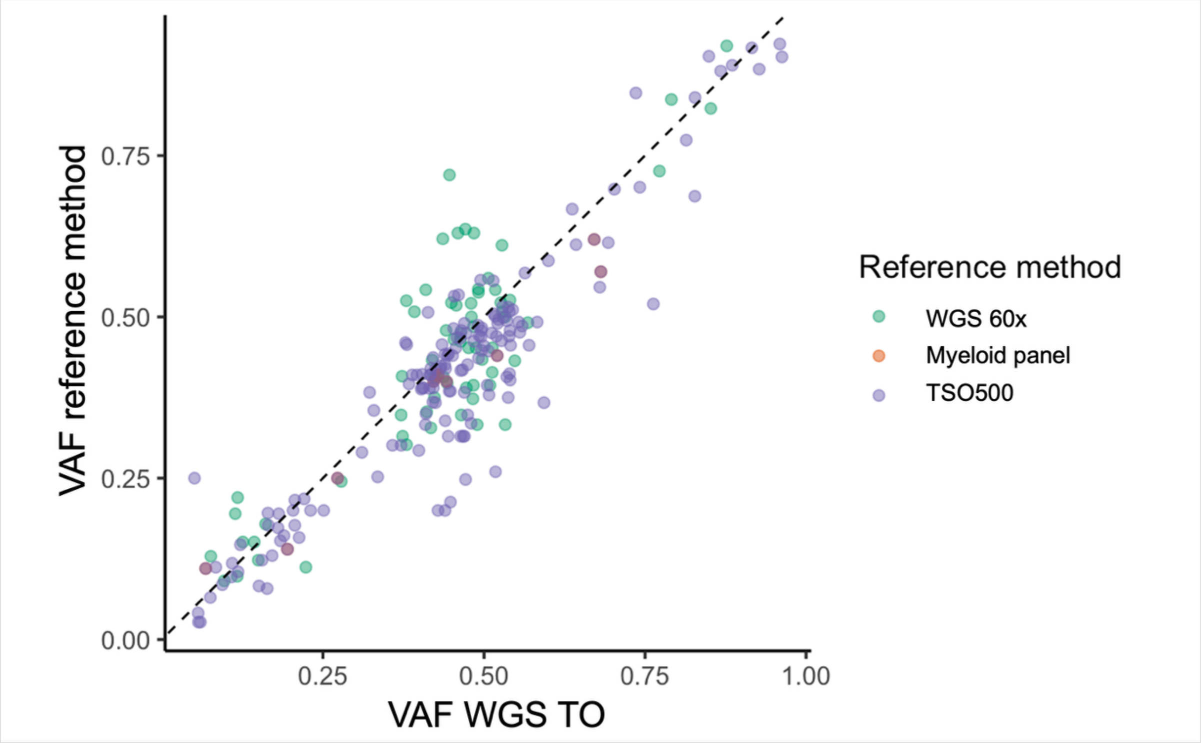

**Supplemental Figure 2. Comparison of VAF in WGS-TO and reference set.** The correlation between the VAF values of the orthogonal set (y-axis) and the WGS TO assay (x-axis). Points are color-coded to indicate variants from different orthogonal methods.

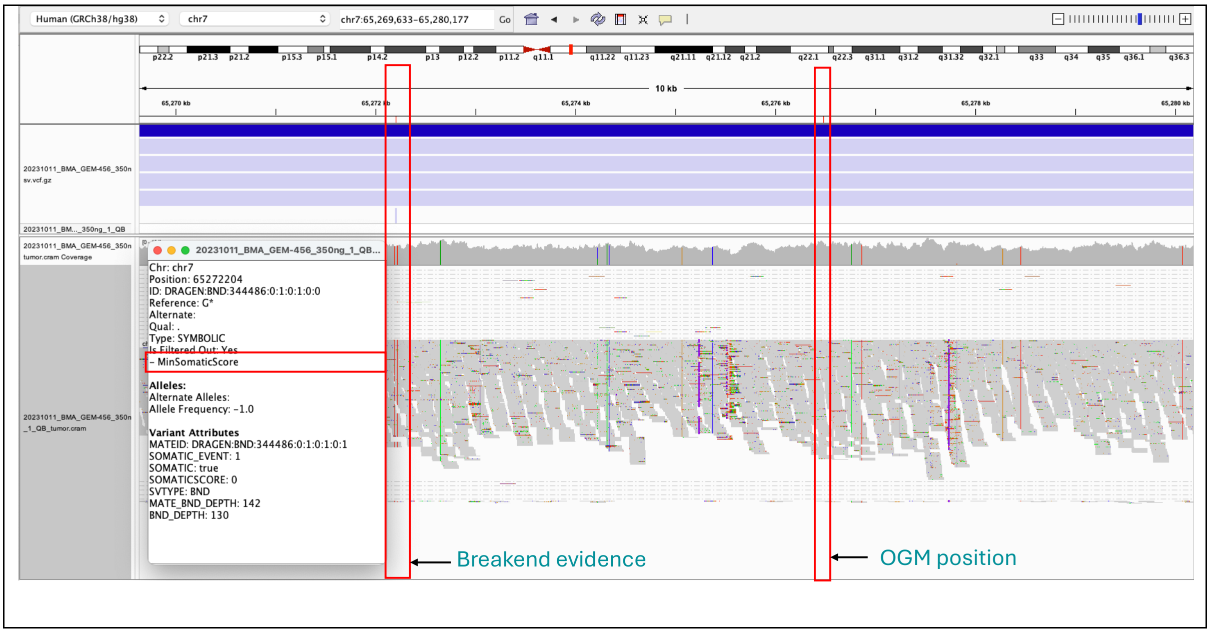

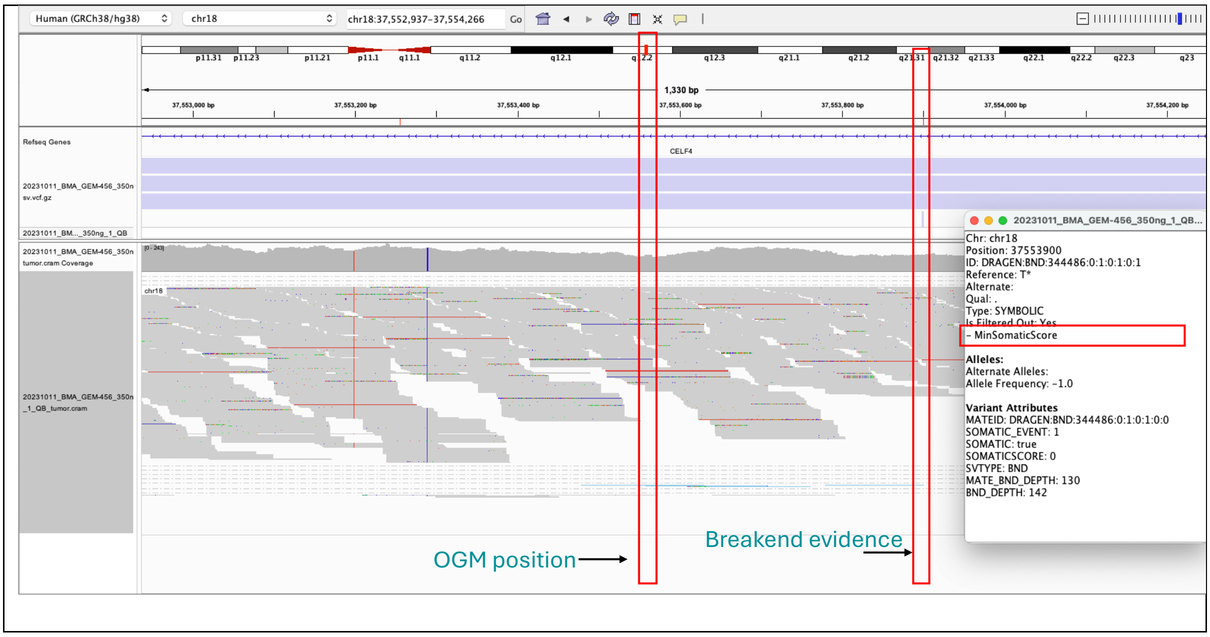

**Supplemental Figure 3. IGV Investigation of missed SV: t(7;18)(q11.21;q12.2)(65276501;37553275).**

In CohortC_456, this variant was reported in OGM and not in WGS TO. The top and bottom panels report the genomics regions around the breakends reported in the orthogonal set (chr7:65276501and chr18: 37553275). In the same cytobands, DRAGEN identified evidence of breakends, but these are not PASS.

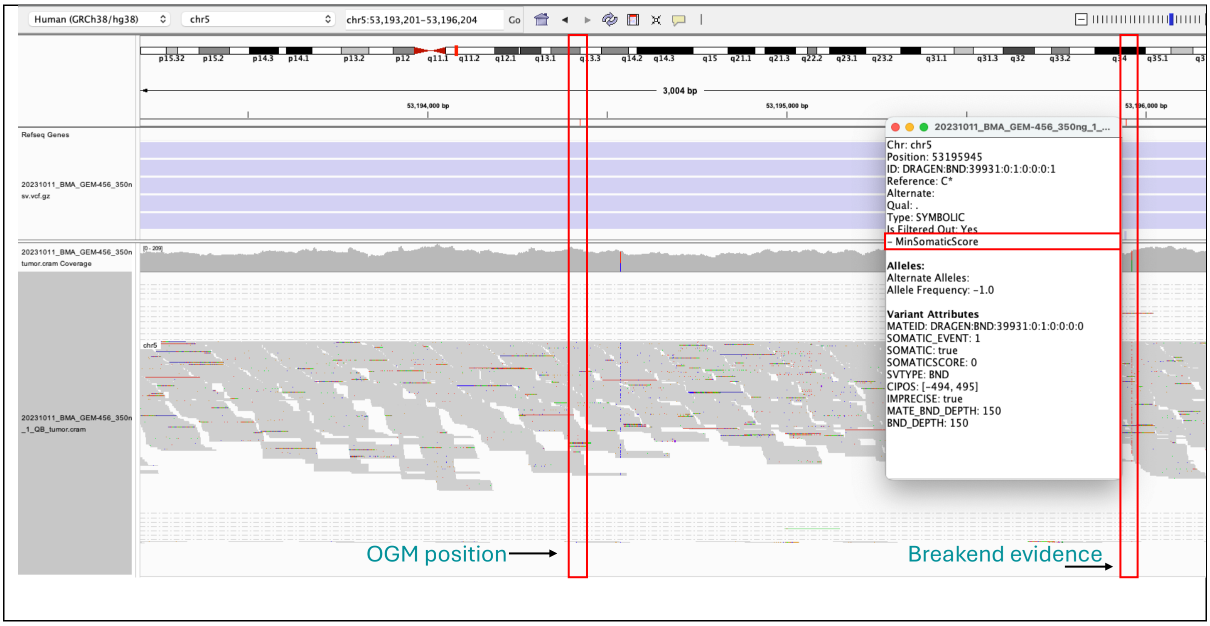

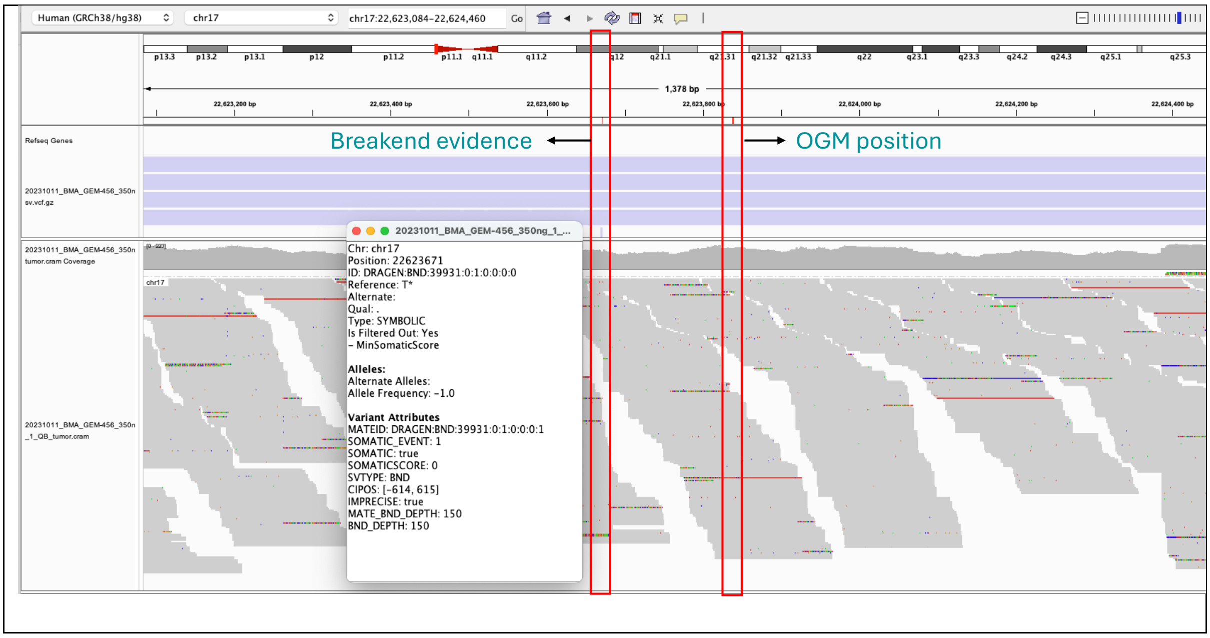

**Supplemental Figure 4. IGV Investigation of missed SV: t(5;17)(q11.2;p11.2)(53194444;22623858).**

In CohortC_456, this variant was reported in OGM and not in WGS TO. The top and bottom panels report the genomics regions around the breakends reported in the orthogonal set chr5: 53194444chr17: 22623858). In the same cytobands, DRAGEN identified evidence of breakends, but these are not PASS.

#### Supplemental Tables

**Supplemental Table 1. List of variants examined for sensitivity.**

Variants are grouped by variant type and orthogonal testing methods.

**Supplemental Table 2. Experiment set up for LoD and list of variants examined for LoD**.

Seracare was titrated to create a series of samples with different target tumor fraction and down-sampled at multiple coverage level to establish LoD. Listed are the number of replicates at each target tumor fraction and coverage level.

**Supplemental Table 3. List of missed variants**.

Variants are grouped by cohort.

**Supplemental Table 4. List of variants examined for analytical precision.**

**Supplemental Table 5. Additional variants in WGS-TO assay compared to orthogonal methods.**
